# Assessing Episodic and Semantic Autobiographical Recall in Healthy Older *APOE* ε4 Carriers

**DOI:** 10.1101/2025.07.20.25331700

**Authors:** Riccardo Sacripante, Tabitha James, Greta Melega, Fiona Lancelotte, Ann-Kathrin Johnen, Cristian Lopez Saquisili, Andreas Lidström, Lingzi Niu, Kayleigh Goddard, Samuel J. Fountain, Ian Clark, Joshua Blake, Michael Hornberger, Brian Levine, Louis Renoult

**Author notes:** **Corresponding author:** Norwich Medical School, University of East Anglia, Norwich Research Park, Norwich, NR4 7TJ, UK. **Author’s note:** This study was supported by a Medical Research Council grant (MRC-MR/S011463/1) allocated to Dr. Louis Renoult and was completed as part of a thesis portfolio for the Doctorate of Clinical Psychology at the University of East Anglia of Riccardo Sacripante. The authors thank all participants who took part in our study, and Lisa Poberezhniuk and Josephine Owusuwaah for their help with the transcriptions of the interview. Scored data and additional online materials are available at the project’s Open Science Framework (OSF) page at https://osf.io/ewdzu/?view_only=8c57930ba2754594a0c6968c91403128 (raw data cannot be provided as they cannot be fully anonymised). For open access, the author has applied a CC BY public copyright license to any Author Accepted Manuscript version arising from this submission. Portions of these data were presented as a talk at the 2025 Cambridge Memory Meeting (University of Cambridge, UK) and at the 2025 Spring Meeting of the British Neuropsychological Society (University College London, UK). Riccardo Sacripante covered a lead role in the data curation, formal analysis, visualization, and writing the original draft of the paper. Riccardo Sacripante, Louis Renoult, Tabitha James, Greta Melega, Fiona Lancelotte, Ann-Kathrin Johnen, and Brian Levine played a role in the conceptualization of the study. Riccardo Sacripante, Tabitha James, Greta Melega, Fiona Lancelotte, Ann-Kathrin Johnen, Cristian Saquisili Lopez, Andreas Lidström, Lingzi Niu and Kayleigh Goddard contributed to the investigation of the study. Louis Renoult, Samuel Fountain, Ian Clark, Joshua Blake, and Michael Hornberger worked on the project administration. Louis Renoult played a central role in funding acquisition, while Joshua Blake and Louis Renoult provided supervision of this study. All the authors reviewed and edited the final draft before submission.

## Abstract

The *APOE* ε4 gene is associated with increased risk of developing sporadic Alzheimer’s Disease (AD). Several studies have focused on declarative memory, where episodic memory deficits are reported in ε4 carriers, while semantic memory has received much less attention. To clarify whether the impact of *APOE* ε4 on declarative memory is specific to episodic memory, we administered a novel measure of autobiographical memory, the Semantic Autobiographical Interview (SAI). Thirty-eight healthy older adults were recruited, 19 ε4 carriers and 19 non-carriers, matched in age, education, and gender. The groups did not significantly differ in any neuropsychological tests except for recognition memory, where ε4 carriers showed reduced performance. On the original Autobiographical Interview (AI), results revealed a reduced number of episodic details (internal details and external events) in carriers. Together, these results suggest a reduction of episodic specificity in ε4 carriers. In contrast, carriers had very similar semantic production to non-carriers, whether it was for off-task (external) semantic details in the AI, or on-task general and personal semantic details produced in the SAI. These results suggest that older adults retain the gist of their personal experience and that the semanticization of their autobiographical narratives is robust and less sensitive to risk for AD than episodic memory.

**Public Significance Statement:** People at risk for Alzheimer’s Disease struggle with recalling memories from specific events (episodic memory), but it remains unclear whether they also have difficulties with personal and general knowledge (semantic memory). We used a new measure, the Semantic Autobiographical Interview, to evaluate these forms of memory. Compared to control participants, people at risk for Alzheimer’s Disease produced a reduced number of episodic details, but a similar amount of semantic knowledge.

## Introduction

Narrative analysis provides ecologically valid measures of naturalistic autobiographical memory recall that are sensitive to subtle memory impairments (Irish, 2023; Simpson et al., 2023), especially when cognitive deficits evade standardized neuropsychological assessment (see Fan et al., 2024). To evaluate autobiographical recall specifically in healthy cognitive aging, Levine and colleagues (2002) designed an interview protocol that distinguish between episodic and semantic memory details during event narration, the Autobiographical Interview (AI). In the AI, participants are instructed to retrieve specific personal memories (i.e., from a specific time and place) from different life periods through a graded retrieval procedure starting from free recall and followed by general and specific probes. Levine et al. (2002) reported that older adults recalled fewer episodic (or internal) details related to the events than younger adults, but also showed a higher recall of external details (i.e., semantic details, unsolicited repetitions, comments, metacognitive statements, and details about events not related to the main episode selected by the participant) across the life periods selected. The authors, therefore, concluded that episodic memory is somehow affected in healthy aging, while semantic memory seems to be better preserved, or even facilitated (see also St Jacques & Levine, 2007). Several explanations have been advanced to account for the increase in external details in aging: older adults could show difficulties with suppressing non-pertinent information during recall, likely due to impaired executive control (Amer et al., 2016, 2018, 2019, 2022; Levine, 2004; Spreng et al., 2018), while others argued that this enriched production of external details may act as compensation for reduced production of internal details (Devitt et al., 2017; Sheldon et al., 2024; for a review, see Grilli & Sheldon, 2022).

Ever since this initial study, the AI has been employed in hundreds of studies involving clinical and healthy cohorts. In a recent meta-analysis, Simpson et al. (2023) noted that the finding of reduced internal details and increased external details in healthy aging is evident and robust across nearly all the reviewed studies. Moreover, compared to healthy aging, the retrieval of episodic internal details on the AI is increasingly more affected in Mild Cognitive Impairment (MCI) and, to a greater extent, in Alzheimer’s Disease (AD), while the effect of increased recall of external details gradually fades with the presence of MCI and AD.

Similar results have been observed using other tests of autobiographical memory in healthy aging, such as the Autobiographical Memory Test (AMT), where older adults typically retrieve a smaller amount of specific memories (e.g., Barry et al., 2020; Ros et al., 2009; 2018), the TEMPau (Piolino et al., 2003, 2006) where a reduction in sensory-perceptive, affective or spatiotemporal details has been observed, or the Episodic Autobiographical Memory Interview (EAMI; Irish et al., 2011), where older adults exhibited a recency effect with progressively increased recall of contextual details for more recent life periods.

While this research has provided extensive insights into episodic memory recall in aging, the role of semantic memory remains to be clarified. External details include metacognitive statements, comments and repetitions, and details about off-topic events, and thus do not constitute a pure measure of semantic processing. Moreover, the instructions of the original AI (and of most tests of autobiographical memory) are to recall episodic memories. Semantic details are incidental and thus do not represent a true assessment of semantic memory, as illustrated by the fact that they are sparsest in healthy younger participants, who presumably have no semantic processing impairment (Renoult et al., 2020; Simpson et al., 2023). Finally, the category of semantic details as originally defined did not distinguish between personal semantics that are specific to the individual (e.g., someone’s favourite movie or knowledge about one’s preferences) from general semantics that are culturally shared and pertain to public and accessible information (e.g., the name of a Prime Minister during a specific historical period). These distinct categories within semantic memory are differentially affected by aging and neurodegenerative disease (Acevedo-Molina et al., 2020; Melega et al., 2024; Renoult et al., 2020).

### Personal and General Semantics: the Semantic Autobiographical Interview

Renoult et al. (2020) introduced a new scoring procedure for the AI to differentiate subtypes of external details and test whether the elevation of these details in aging, in frontotemporal lobar degeneration (including mixed frontotemporal/semantic dementia:FTD/SD), and in progressive non-fluent aphasia was specific to general and personal semantics or concerned all subtypes. While the increase in external details concerned all types of semantic details (both personal and general) in healthy older adults (see also Acevedo-Molina et al., 2020), participants with FTD and SD showed an excess of personal semantic but not general semantic details. These results can be related to the observations that patients with FTD – in particular SD – have an impairment in recalling general semantic knowledge (Lambon Ralph et al., 2017), but better preservation of personally relevant concepts (Duval et al., 2012; McKinnon et al., 2008).

Melega and colleagues (2024) recently developed the Semantic Autobiographical Interview (SAI) that directly targets personal and general semantics in different sections of the interview, allowing a less ambiguous interpretation of semantic recall in aging. The SAI was tested alongside the original AI among healthy older and younger adults, and it was found that older adults reported lower proportions of target details (i.e., details probed by instructions) and more external details not probed by the instructions across the three interviews (i.e., off-task utterances or ‘stories aside’; see Bluck et al., 2016). As compared to young adults, older adults also consistently produced more autobiographical facts and self-knowledge across interviews, which could reflect a bias towards personal semantic information in healthy aging regardless of task instructions. Although the study also observed the typical reduction of internal details in older adults in the AI, the other findings suggest that the shift in narrative style among older adults goes beyond episodic remembering (Bluck et al., 2016; Hasher & Zacks, 1998; James et al., 1998; Trunk & Abrams, 2009).

### Autobiographical Memory and *APOE* **ε**4

Given the potential of autobiographical memory retrieval as a sensitive and subtle measure of cognitive decline in people at the preclinical stage of AD, recent research has extended these investigations to healthy people at increased genetic risk of developing the disease. The ε4 variant of the Apolipoprotein (*APOE*) gene is associated with an increased risk of developing sporadic late-onset Alzheimer’s Disease (AD) (Borgaonkar et al., 1993; Corder et al., 1993), with an earlier age of onset (Blacker et al., 1997). Since *APOE* ε4 is the strongest genetic risk factor for late-onset AD (Fortea et al., 2024), several studies have focused their attention on declarative memory, where episodic memory deficits have been reported in ε4 carriers (O’Donoughe et al., 2018; Small et al., 2004; Wisdom et al., 2011), consistently with its early impairment in AD (McKhann et al., 2011). In contrast, the impact of *APOE* ε4 on semantic memory has received much less attention, though it has also sometimes been reported to be impaired in mild cognitive impairment and early AD (Chasles et al., 2020; Joubert et al., 2010, 2020; Storandt, 2008; Taler et al., 2016, 2020).

A study using the AI in ε4 carriers reported a reduced number of internal episodic details during autobiographical recall, but no difference in external details relative to non-carriers (Grilli et al., 2018), suggesting a selective impairment of episodic memory in ε4 carriers. Similar findings in *APOE* ε4 carriers were also observed by Acevedo-Molina et al. (2023) when assessing past and future autobiographical thinking. However, as stated above, external details in the AI not only include general and personal semantic details, but also metacognitive statements, comments and repetitions, and details about off-topic events, and are thus not a pure measure of semantic processing. In a related study (Grilli et al., 2021), older *APOE* ε4 carriers showed a deficit in the generation of episodic and personal semantic memories on an autobiographical memory fluency task (Addis & Tippett, 2004; Dritschel et al., 1992). More recently, Knoff et al. (2024) reported that *APOE* ε4 carriers could also show an increased difficulty in the rapid reconstruction of specific autobiographical memories without the aid of semantic memory. However, as mentioned above, most tests of autobiographical memory instruct participants to retrieve episodic memories, and thus the interpretation of semantic details (and external details in the AI) remains ambiguous.. What is therefore still missing in current research is a direct comparison of episodic memory and personal and general forms of semantic memory when assessing autobiographical recall.

### The present study

To address this issue and clarify whether the impact of *APOE* ε4 on declarative memory is specific to episodic memory, in the present study we administered a new measure of autobiographical memory, the SAI (Melega et al., 2024), to cognitively unimpaired older *APOE* ε4 carriers and non-carriers, alongside the original AI. Such comparison could cast light on the function of personal and general semantic details among healthy older people with increased genetic risk for AD and help verify whether cognitive decline in this population is specific to episodic memory.

In line with previous research (Grilli et al., 2018), we hypothesised that *APOE* ε4 carriers would show decreased episodic recollection of event-specific experiences (i.e., episodic details). As to personal and general semantic memory details, it remains unclear whether *APOE* ε4 carriers would show an increased or a decreased recall of these details, given that previous research in the field did not directly probe for semantic details as indicated by the standard AI instructions (Grilli et al., 2018) or used different research paradigms (see Grilli et al., 2021). The existing literature suggests that the impact of *APOE* ε4 on semantic memory may be limited (Buckley et al., 2014; Wilson et al., 2002; for a review see Sacripante et al., 2025), though there are only a few relevant studies, and they mostly used simple neuropsychological tests like naming that are not a good match for episodic tests, as well as tests like category fluency that also rely on executive functions (see Nilsson et al., 2006). It could be speculated that forms of personal semantics such as memories of repeated events, that have similar neural correlates as episodic memory (Renoult et al., 2012), are likely to be more affected in *APOE* ε4 carriers than other forms such as autobiographical facts, that are more similar to general semantics (Grilli et al., 2021), but the evidence so far remains limited.

## Methods

### Transparency and Openness

Scored data and analysis code are available at https://osf.io/ewdzu/?view_only=8c57930ba2754594a0c6968c91403128 (raw data cannot be released as these data cannot be fully anonymized). This study’s design and its analysis were not pre-registered. Data were analysed using the R Studio statistical software version 4.0.3 (R Core Team, 2020).

### Participants

A total of 38 healthy older adults with an average age of 67.76 years of age (15 women, 23 men, range: 61-83 years, *SD* = 6.05) and an average of 15.31 years of education (range: 6-26 years, *SD* = 3.41) were recruited at the School of Psychology of the University of East Anglia. Participants were contacted by the research team through advertisements to the university’s volunteer panel, as well as a press release and a radio interview.

To be considered eligible for the study, all participants needed to be over 60, to be either English native speakers or having learned English early in life, to have normal or corrected to normal vision (i.e., wearing glasses or contacts, as required) and have no diagnosed psychiatric or neurological conditions.

We conducted an a priori power analysis for a mixed factorial ANOVA using G*Power (Faul et al., 2009), which indicated a minimum total number of 36 participants, assuming a power of 0.90, a medium effect size (η*2_p_* = 0.06, equal to an effect size of 0.25) and an alpha probability of .05. Our sample consisted of 19 ε4 carriers (ε3ε4 n = 16, ε4ε4 n = 2, and ε2ε4 n = 1) and 19 non-carriers (ε3ε3 n = 16, and ε2ε3 n = 3). The two groups were similar for age, years of education and gender (see *Table 1*). Detailed information about DNA extraction and APOE genotyping are provided in the *Supplementary Material*.

**Table 1.**
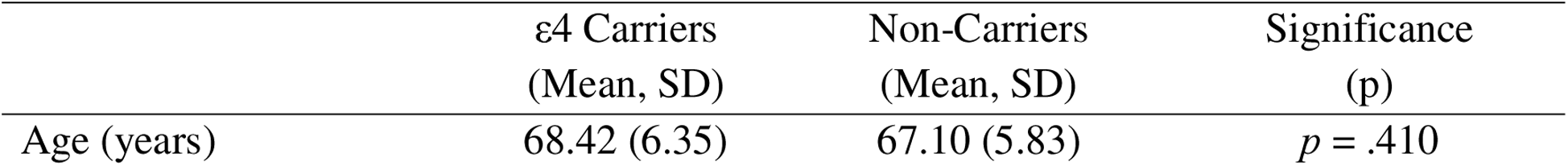
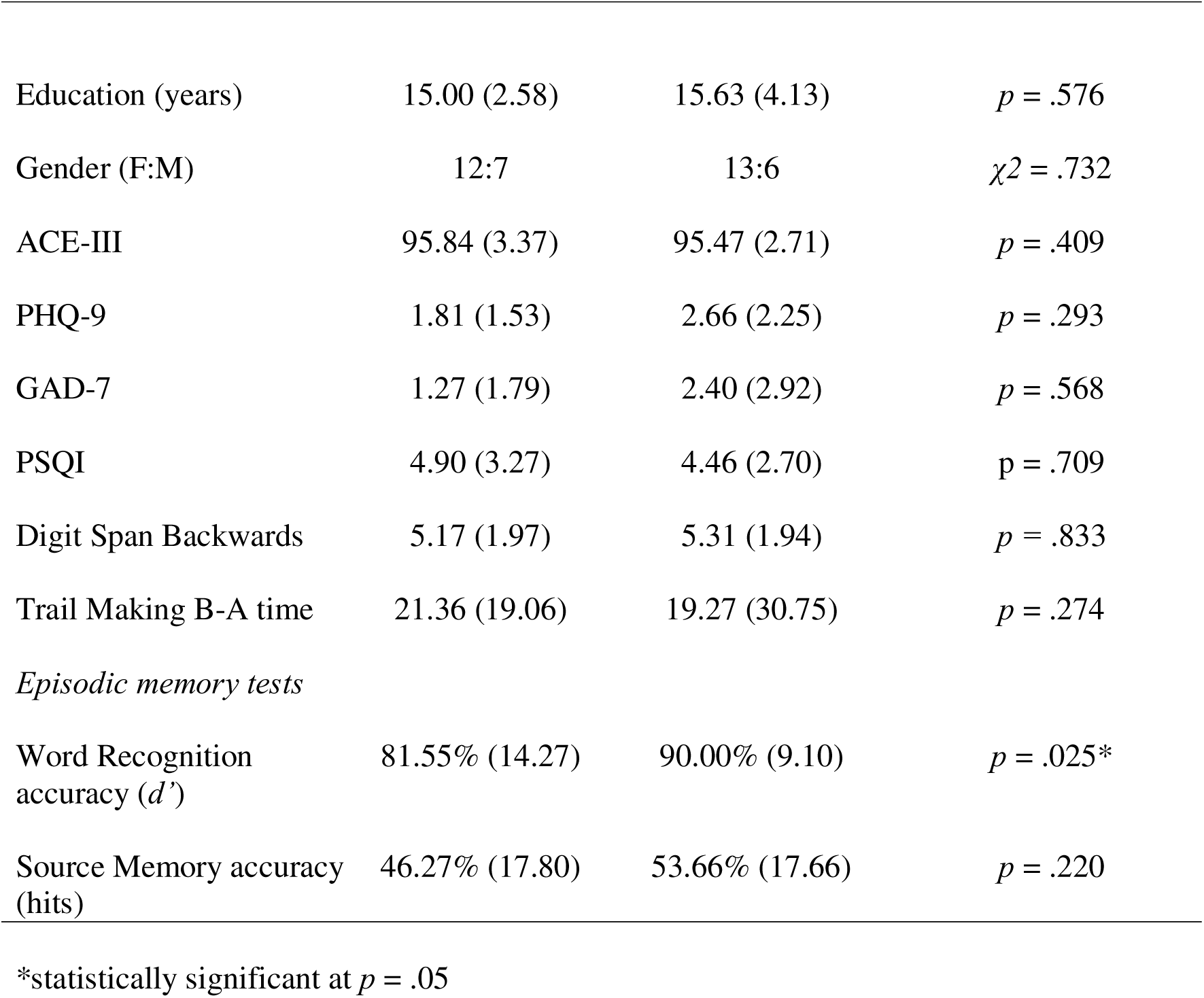
Demographics and mean scores on the questionnaires and neuropsychological tests for ε4 carriers and non-carriers.

| | ε4 Carriers<br>(Mean, SD) | Non-Carriers<br>(Mean, SD) | Significance<br>(p) |
| --- | --- | --- | --- |
| Age (years) | 68.42 (6.35) | 67.10 (5.83) | $p = .410$ |
| Education (years) | 15.00 (2.58) | 15.63 (4.13) | $p = .576$ |
| Gender (F:M) | 12:7 | 13:6 | $\chi^2 = .732$ |
| ACE-III | 95.84 (3.37) | 95.47 (2.71) | $p = .409$ |
| PHQ-9 | 1.81 (1.53) | 2.66 (2.25) | $p = .293$ |
| GAD-7 | 1.27 (1.79) | 2.40 (2.92) | $p = .568$ |
| PSQI | 4.90 (3.27) | 4.46 (2.70) | $p = .709$ |
| Digit Span Backwards | 5.17 (1.97) | 5.31 (1.94) | $p = .833$ |
| Trail Making B-A time | 21.36 (19.06) | 19.27 (30.75) | $p = .274$ |
| *Episodic memory tests* |  |  |  |
| Word Recognition accuracy ($d'$) | 81.55% (14.27) | 90.00% (9.10) | $p = .025^*$ |
| Source Memory accuracy (hits) | 46.27% (17.80) | 53.66% (17.66) | $p = .220$ |

\*statistically significant at $p = .05$

As part of the recruitment process, participants were screened to exclude any neurological, psychiatric, and other medical conditions that could affect their memory performance. To rule out any cognitive decline, participants completed the Addenbrookes Cognitive Examination (ACE-III; Hsieh et al., 2013; Mioshi et al., 2006) as a screening tool with a cut-off score of 88, and an extended neuropsychological assessment for global cognition through the online platform NeurOn (https://neuropsychology.online/). The battery included the Digit Span Backwards (Weschler, 1987), the Trail Making Test Part A and Part B (Reitan, 1958), and tests of word recognition and source memory (for more details on these tasks, see Melega et al., 2024).

To control any potential group differences that could have influenced or accounted for different patterns of memory recall, all the participants were also asked to complete screening for anxiety (Generalised Anxiety Disorder, GAD-7; Spitzer et al., 2006), depression (Patient Health Questionnaire, PHQ-9; Kroenke et al., 2001) and quality of sleep (Pittsburgh Sleep Quality Index, PSQI; Buysse et al., 1989). These questionnaires were completed through Qualtrics (Qualtrics International, Inc., Provo).

This research project was granted ethical approval from the Research Ethics Committee of the School of Psychology of UEA and the National Research Ethics Service (NRES) for the dementia Research and Care Clinical protocol. All the participants provided their informed consent before taking part in the study and they were free to withdraw at any time. Participants received an honorarium for their research participation. Data was collected online between February 2022 and March 2024.

### Experimental Procedure

The experimental procedure included multiple online sessions via Microsoft Teams. Ahead of testing, participants were asked to provide demographic information and complete the psychological questionnaires (GAD-7, PHQ-9, and PSQI) online through Qualtrics.

The first online session took approximately one hour, and it involved the completion of cognitive screening (ACE-III, online version) and neuropsychological tests for executive functions (Digit Span Backwards, Trail Making Test A and B), and episodic memory (tests of word recognition and source memory) using the online platform NeurOn (https://neuropsychology.online/). A total of 35 participants completed the battery of neuropsychological tests. One carrier failed to complete the digit span backward only, another carrier did not complete both the digit span backward and the episodic memory tests, and one non-carrier participant did not complete any of these tests.

After completing these tests, in preparation for the autobiographical interview, participants were asked to list personal chapters from their lives by segmenting their last 30 years as chapters. Every participant listed as many chapters as they wished, whereby each chapter ranged from one to five years long. Every listed chapter was assigned a title with beginning and ending dates.

The second session took place online via Teams between two and seven days after the initial one, and it took a minimum of one hour to be completed, with breaks provided to avoid participants’ fatigue. Participants were asked to consider the life chapters from last year and from ten years ago, which were used as personalised temporal cues for memories throughout the different sections of the interview. If participants had provided multiple chapters from the same years (e.g., ten years ago), they were then asked to choose the chapter they felt most confident with. Crucially, both life chapters were recalled in the AI and the personal semantic version of the SAI (i.e., PSAI), while the general semantic version of the SAI (i.e., GSAI) only concerned the most recent (i.e., last year), as in Melega et al. (2024)^1^.

As the transcripts represented the main source of data for this study, the whole online session was recorded and transcribed simultaneously. Participants completed the original AI, the PSAI and the GSAI. On the AI, participants were asked to describe in detail a specific event from two life chapters selected (i.e., last year and ten years before), with general probes (i.e., follow-up prompts to encourage further elaboration or to maximise the retrieval of episodic details), and specific probes regarding spatiotemporal, perceptual, and emotional aspects concerning the selected event in line with the administration manual of the AI (see Levine et al., 2002). While undertaking the PSAI, participants were instructed to describe what was going on in their life during the selected life chapter (instruction: “*if you wanted to tell me how that life chapter was like for you, how would you describe it?*”, see Melega et al., 2024). On the PSAI, specific probes concerned autobiographical facts (regarding personally relevant facts, people and places), repeated events (e.g., weekly habits, routines, hobbies), and self-knowledge information (e.g., personality traits and character, opinions, and beliefs, preferences) from the same two life chapters as in the AI (i.e., last year and ten years before). The order of administration of the original AI and the PSAI was counterbalanced for each participant.

After the AI and the PSAI, all participants completed the GSAI, where they were asked to recall general semantic information, defined as culturally shared general knowledge (Renoult & Rugg, 2020; Renoult et al., 2020). Thus, participants had to describe what was going on in their local community, country, or around the world (instructions: “*if you wanted to tell me what was happening in your community, in the UK or around the world during that specific life chapter, what would you say?*”). On the GSAI, specific probes concerned public events, public figures, trends and popular things (e.g., films, music, fashion). An illustration of the experimental paradigm can be found in *Figure 1*.

**Figure 1.**
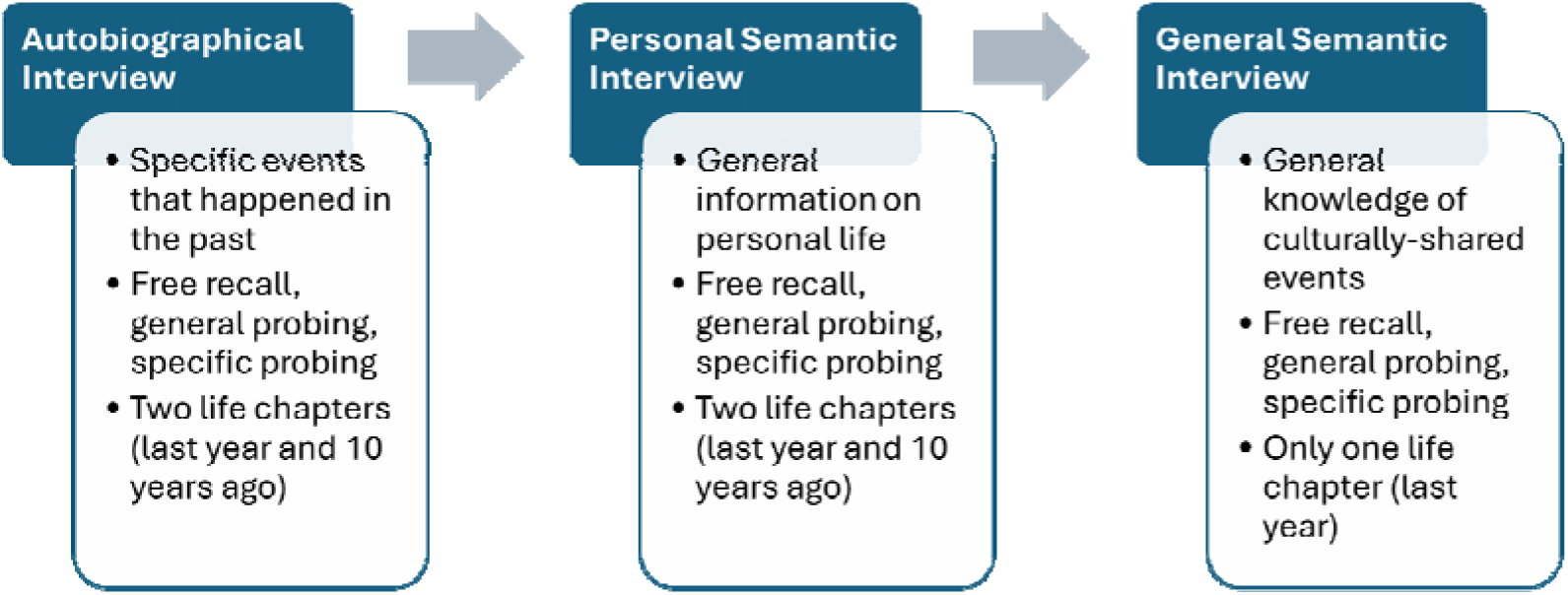
Illustration of the experimental paradigm involving the three online interviews (AI, PSAI, and GSAI). The order of administration of the AI and the PSAI was counterbalanced.

### Detail scoring procedure

As stated above, all participants consented with the interviews being recorded and automatically transcribed using Microsoft Teams and then manually edited by three researchers (RS, TJ, GM). In line with Melega et al. (2024), memories were scored following the method described by Levine et al. (2002) and the recent taxonomy of semantic details of Renoult et al. (2020). Interview transcripts were segmented into different types of details which were classified as episodic, autobiographical facts, self-knowledge, repeated events, general semantic, repetitions, and other. *Table 2* provides definitions and examples for each type of detail.

**Table 2.** Definition of Detail types and relative examples.

| Detail Type | Definition | Examples |
| --- | --- | --- |
| Episodic | Unfolding of the event, spatiotemporal, perceptual and emotional details. | Last year I visited Mexico; I was quite fascinated; There was a crocodile on the right side of the riverbank. |
| Autobiographical fact | Basic (objective) information about personal life circumstances, factual element of unique episodes. | I lived in Scotland for a few years; I have a younger sister. |
| Self-knowledge | Personality traits and character, opinions and beliefs. | I was very happy during that time; I quite like watching football. |
| Repeated Event | Common elements of repeated episodes. | I go to the gym three times a week; I walk to the office every day. |
| General Semantic | Culturally shared knowledge (e.g., neighbourhood community, country, world). | Rome is the capital of Italy; the UK general elections took place this year. |
| Repetition | Information that has already been recalled. | As I mentioned, the UK general elections happened this year. |
| Other | Metacognitive statements and editorializing. | Right, let me have a think; I don't know; I can't think of anything that happened during that time. |

Interview transcripts were rated by two independent scorers (RS, TJ) who were trained on the original AI scoring method (Levine et al., 2002), as well as on the novel scoring method for external details (Renoult et al., 2020). To estimate interrater reliability, six memories from the original AI (15.8%), six from the PSAI (15.8%), and six from the GSAI (15.8%) were randomly selected and scored by both scorers, who were blind to the group allocation of each participant. As in Melega et al. (2024), we calculated interrater reliability separately for each interview by referring to Intraclass Correlation Coefficient (ICC; two-way, random effects model). After collapsing the specific categories into macro-categories (Internal vs External), inter-rater reliability on internal details was excellent across interviews (ICCs for the AI > 0.96, ICCs for the PSAI > 0.92, ICCs for the GSAI > 0.97), as well as for external details (ICCs for the AI > 0.92, ICCs for the PSAI > 0.85, ICCs for the GSAI > 0.97).

The interview transcripts from the remaining participants were allocated to the scorers in pseudorandom order, by making sure they were blinded to the group allocation of each participant scored (i.e., ε4 carriers or non-carriers), which could have biased their scoring.

We used a script developed in MATLAB (Mathworks, Inc.) for automated details counting on the interview transcript of each participant, to ease scoring and to minimise human error (see Wardell et al., 2021; Melega et al., 2024).

### Data Processing and Analysis Plan

Ahead of data analysis, we employed a winsorization process for all the scored memories to re-adjust positively skewed data (see Melega et al., 2024; Renoult et al., 2020). Winsorization is a statistical method used to reduce the influence of outliers in datasets. Using this procedure, we rescaled detail counts that were ± 2.5 SD from the mean to 2.5 SD from the mean (see McKinnon et al., 2008, 2015), for a total of 43 winsorized data points, which accounted for 3.05% of the total scores (for carriers, 1.31%, 2.85%, 1.42% of AI, PSAI, GSAI, respectively; for non-carriers, 5.60%, 3.21%, 2.14% of AI, PSAI, GSAI scores, respectively). As in Melega et al. (2024), we averaged detail counts across recent and remote memories, since the effect of time-period was not significant.

In this study, we report the analyses on averaged cumulative scores, which include free recall, general probe, and specific probe. This approach allowed us to refer to more robust estimates of group differences due to increased observations and smaller error variance (see Melega et al., 2024). Detail counts of cumulative recall represented our main measure of interest. Although we did not expect differences in narrative length due to verbosity between our two groups of older adults, proportional details (i.e., details count divided by the total number of details produced by each participant) were also considered as a measure of interest for cumulative recall (see *Supplementary material*).

As in Melega et al. (2024), target detail scores corresponded to episodic details for the AI, personal semantic details (autobiographical facts, self-knowledge, and repeated events) for the PSAI, and general semantic details for the GSAI. For the analysis of target details between interviews, we employed a 2×3 mixed design, with Group as a between-subjects variable (carriers vs non-carriers), Interview as within-subjects variable (AI, PSAI and GSAI) with the respective target detail score on each interview as an outcome variable.

To consider participants’ differences in the elaboration of specific details within each interview, we also adopted a 2×7 mixed design, with Group as between-subjects variable, and Detail Type (Episodic, Autobiographical Facts, Self-knowledge, Repeated Events, General Semantic events, Repetition, Other) as within-subjects variable. For the original AI, internal details and external events were grouped into a single cumulative category (Episodic details), for more direct comparison of results with the PSAI and the GSAI, whereby episodic details are considered external events (see Melega et al., 2024).

Analyses were carried out in R Studio (version 4.0.3). As in Melega et al. (2024), statistical analyses involved mixed factorial analyses of variance (ANOVAs) to assess group differences (carriers vs non-carriers) for target details on each of the three interviews, and the analysis of specific details production within each interview. The assumption for normality of distribution for parametric testing was checked and when this assumption was violated, mixed ANOVAs were also computed using permutation tests as a non-parametric statistical method, with a maximum number of 1000 iterations. Post-hoc tests involving Detail Type and Group were computed via pairwise comparisons of estimated marginal means with emmeans package in R (Lenth et al., 2014) corrected for false discovery rate (Benjamini & Hochberg, 1995).

## Results

No significant group differences were noted between *APOE* ε4 carriers and non-carriers on any of the demographic measures, cognitive screening, and neuropsychological tests, apart from the Word Recognition accuracy index (*d’)* of the episodic memory tests, where carriers performed significantly worse (*p* = .025).

In the overall sample of participants including both carriers and non-carriers, Word Recognition Accuracy positively correlated to the total count of episodic details in the AI (τ = 0.269, *p* = 0.036). After dividing the two groups, the correlations were however non-significant for both carriers (*p* = 0.78) and non-carriers (*p* = 0.48).

### *APOE* group differences in the production of target details across interviews

*Figure 2* shows the total average counts of target details produced by carriers and non-carriers in each interview (see also *Table 3)*. Target details were the composite sum of internal details and external events (Episodic details) in the AI, the sum of Autobiographical Facts, Self-Knowledge, and Repeated Events details for the PSAI, and General Semantic details for the GSAI. This allowed us to make our data more comparable across the three interviews, as in Melega et al. (2024). Statistical analyses on target details with proportional scores can also be found in the *Supplementary Material*.

**Figure 2.**
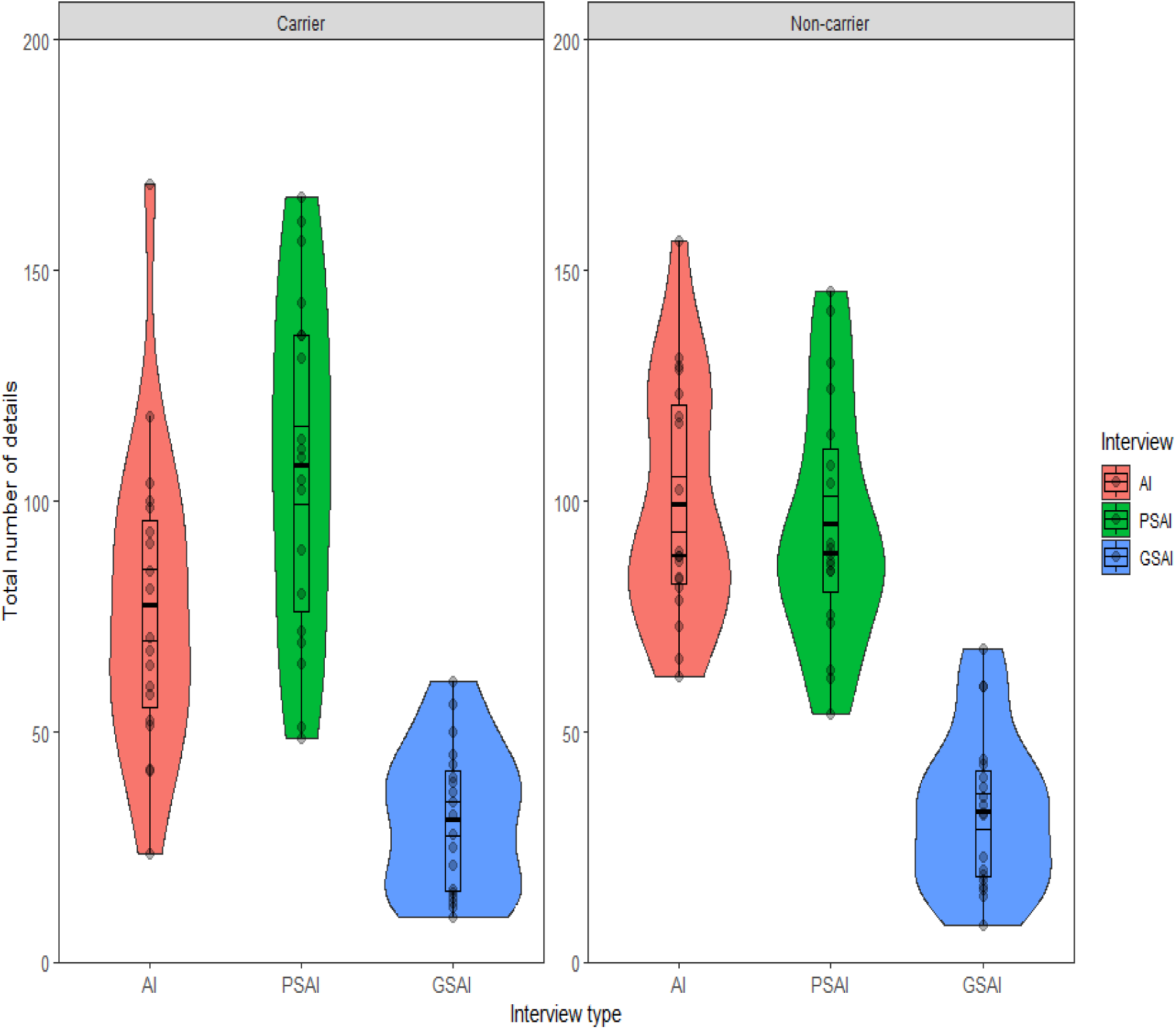
Counts of target details during cumulative recall in the carrier and non-carrier groups across interviews with medians and interquartile range. Target details correspond to the information that was probed by task instructions: episodic details in the Autobiographical Interview (AI); personal semantic details (autobiographical facts, self-knowledge, and repeated events) in the Personal Semantic Interview (PSAI); general semantic details in the General Semantic Interview (GSAI).

**Table 3.**
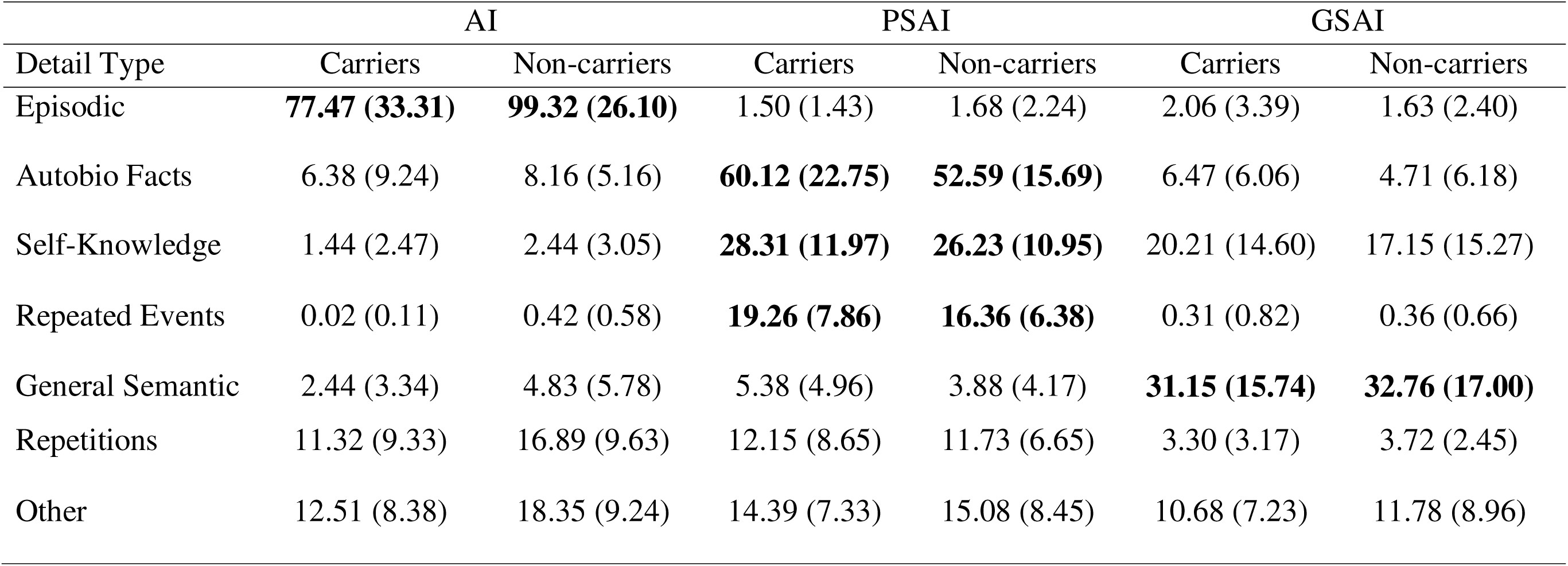
Sum of count scores in carriers and non-carriers for cumulative recall (Free Recall, General Probe, and Specific Probe) in the AI, PSAI, and GSAI.

As illustrated by the main effect of Interview type, *F*(2,108) = 70.996, η*2p* = 0.57, 95% CI [0.47, 1.00], *p* < .001, participants produced a significantly higher amount of target details on the PSAI (*M* = 101.44, *SD* = 32.2) as compared to the GSAI (*M* = 32.0, *SD* = 16.2), *t*(108) = 11.210, *p* < .001, but not compared to the original AI (*M* = 88.40, *SD* = 31.52), *t*(108) = 2.104, *p* = .093. This finding is consistent with Melega et al. (2024), who also reported less target details in the GSAI compared to the PSAI.

There was a significant interaction between Interview and Group, *F*(2,108) = 3.883, η*2p* = 0.07, 95% CI [0.01, 1.00], *p* = .023, as non-carriers produced significantly more target details (*M* = 99.32, *SD* = 26.10) on the original AI than carriers (*M* = 77.47, SD = 33.31), *t*(108) = 2.493, p = .014, while both groups produced similar rates of target details on the PSAI, *t*(108) = -1.427, *p* = .156, as well as on the GSAI, *t*(108) = 0.183, *p* = .855.

When considering target details within each group, carriers produced more target details on the PSAI (*M* = 107.70, *SD* = 36.85) as compared to the AI (*M* = 77.47, *SD* = 33.31), *t*(108) = 3.44, *p* < .01, while this trend was not observed within the non-carriers group, who produced a similar rate of target details on the AI (*M* = 99.32, *SD* = 26.10) and PSAI (*M* = 95.18, *SD* = 26.33), *t*(108) = 0.472, *p* = .884.

### *APOE* group differences in detail elaboration in each interview

To provide a fine-grain analysis around the production of detail categories across the three interviews, we also report data of detail elaboration for each interview (see *Figure 3*). Average count scores on each detail category can be found in *Table 3*. The same detailed analyses were carried out for proportional scores (see *Supplementary Material*).

**Figure 3.**
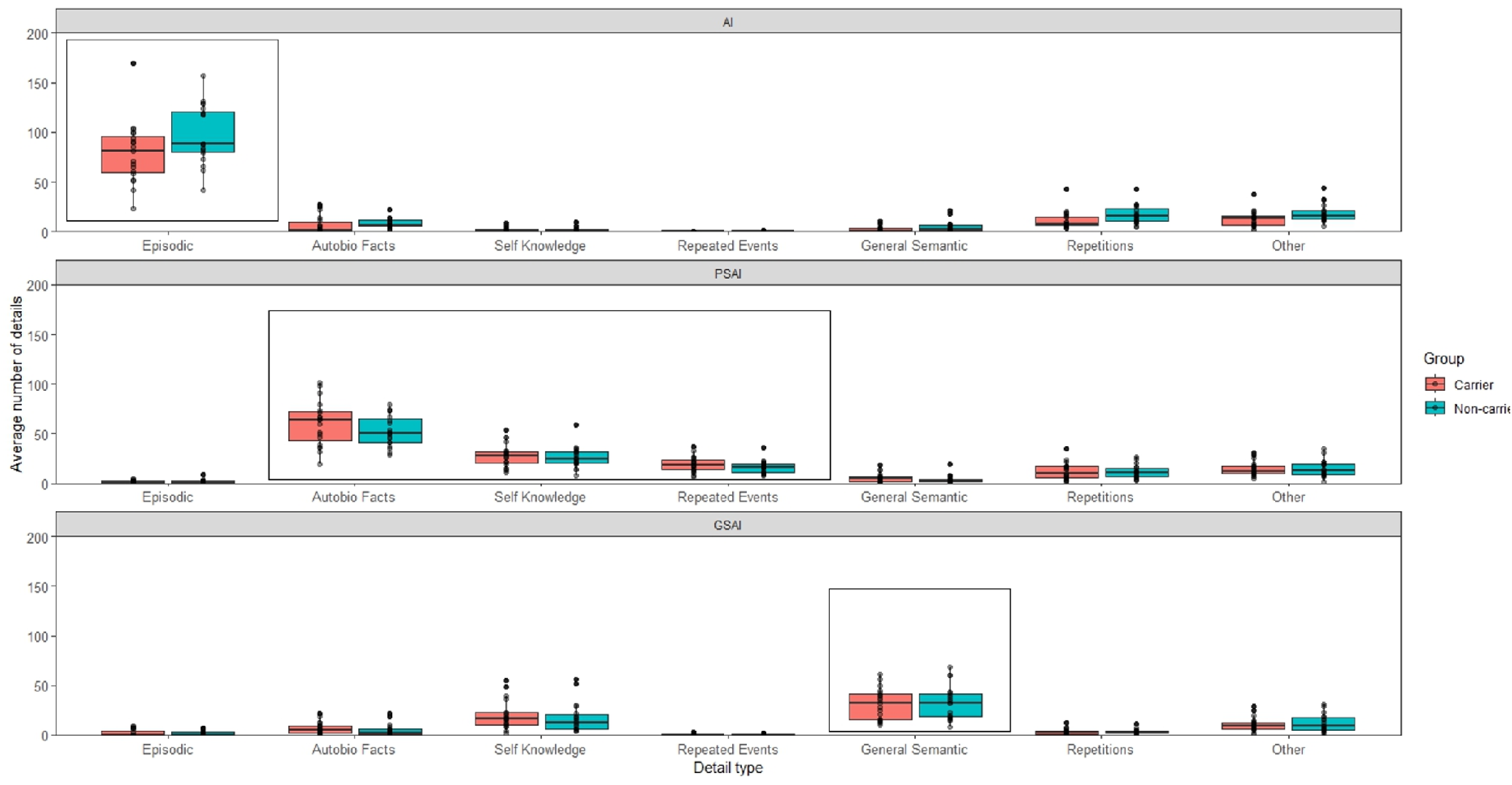
Boxplots showing counts of detail types during cumulative recall in the Carrier and Non-carrier group in the original Autobiographical Interview (AI), Personal Semantic Interview (PSAI), and General Semantic Interview (GSAI). Target details in each interview are highlighted by the black rectangle.

### Autobiographical Interview

The average number of details produced by carriers and non-carriers on the AI is illustrated in *Figure 3* (top panel). ANOVA indicated a main effect of Detail Type, *F*(6,252) = 226.164, η*2_p_* = 0.84, 95% CI [0.82, 1.00], *p* < .001, and a significant main effect of Group, *F*(1, 252) = 12.75, η*2_p_* = 0.05, 95% CI [0.01, 1.00], *p* < .001, where non-carriers overall produced a higher amount of details (*M* = 21.55, *SD* = 34.40) as compared to carriers (*M* = 15.94, *SD* = 29.02), *t*(252) = 3. 571, *p* < .001.

The interaction between Detail Type and Group was also significant, *F*(6,252) = 3.22, η*2_p_* = 0.07, 95% CI [0.01, 1.00], *p* < .01. When comparing groups, post-hoc pairwise comparisons revealed that carriers’ narratives included a reduced number of episodic details (*M* = 77.47, *SD* = 33.31) than non-carriers (*M* = 99.32, *SD* = 26.10), *t*(252) = -5.257, *p* < .001. This finding indicates that episodic details appear to be selectively reduced in carriers. No significant between-group difference was observed for the other types of details (Autobiographical Facts, *t*(252) = 0.536, *p* = 0.60; General Semantic, *t*(252) = 0.57 *p* = 0.56; Other, *t*(252) = 1.40, *p* = 0.16; Repeated Events, *t*(252) = 0.097, *p* = 0.92; Repetitions, *t*(252) = 1.34, *p* = 0.18; Self-Knowledge, *t*(252) = 0.240, *p* = 0.81).

### Personal Semantic Interview

When considering the count data of the different subtypes of details in carriers and non-carriers’ narratives in the PSAI (see *Figure 3*, middle panel), the ANOVA revealed a main effect of Detail Type, *F*(6,252) = 127.24, η*2_p_* = 0.75, 95% CI [0.71, 1.00], *p* < .001, a non-significant main effect of Group, *F*(1,252) = 2.440, η*2_p_*= 0.001, 95% CI [0.00, 1.00], *p* = .120, and a non-significant interaction between Detail Type and Group, *F*(6,252) = 0.721, η*2_p_* = 0.02, 95% CI [0.00, 1.00], *p* = .633.

This indicates that, on the PSAI, carriers showed a very similar semantic production to non-carriers for on-task personal knowledge (autobiographical facts^2^, repeated events, and self-knowledge), and that groups did not significantly differ in terms of how they described past life chapters.

### General Semantic Interview

When considering the count data of the different subtypes of details in carriers and non-carriers’ narratives of life chapters in the GSAI (see *Figure 3*, bottom panel), the ANOVA revealed a main effect of Detail Type, *F*(6,252) = 56.31, η*2_p_* = 0.57, 95% CI [0.51, 1.00], *p* < .001, a non-significant main effect of Group, *F*(1,252) = 0.066, η*2_p_* = 0.002, 95% CI [0.00, 1.00], *p* = .797, and a non-significant interaction between Detail Type and Group, *F*(6,252) = 0.289, η*2_p_* = 0.006, 95% CI [0.00, 1.00], *p* = .942. As observed in the PSAI, these results indicate that carriers showed similar production of on-target general semantic details as non-carriers, as they did not significantly differ on how they described their general knowledge of the world.

## Discussion

The analysis of narrative recall is a reliable method to probe and measure naturalistic forms of autobiographical memory, especially when detecting subtle memory impairments, which could evade standard neuropsychological assessment. When considering declarative forms of memory, people at increased genetic risk of developing late-onset AD commonly show deficits in episodic memory recall (Small et al., 2004; Wisdom et al., 2011), while their semantic memory abilities have received substantially less attention. To verify whether the impact of *APOE* ε4 on declarative memory is specific to episodic memory, we administered the SAI (Melega et al., 2024), which uses instruction manipulations to elicit personal and general semantic autobiographical content.

Overall, both ε4 carriers and non-carriers followed the interview instructions and modulated the content of their narratives so that episodic details were the highest on the original AI, while personal and general semantic details were the highest on the PSAI and GSAI respectively. When considering counts of target details across interviews, carriers produced more target details on the PSAI, especially autobiographical facts and self-knowledge, as compared to the AI and GSAI. Non-carriers produced a similar rate of target details on both the AI and PSAI. Moreover, non-carriers produced more elaborated narratives relating to personal past events, as evidenced by a significantly higher amount of target episodic details on the AI than carriers. These findings suggest a shift in narrative style among ε4 carriers, whereby they produce a lower amount of target episodic details in their narratives. This selective reduction of episodic memory ability was also observed in an episodic memory test of word recognition, where carriers showed reduced performance, in line with previous research that reported episodic memory deficits in this population (O’Donoughe et al., 2018; Small et al., 2004; Wisdom et al., 2011). While others have reported a specific reduction in internal details in carriers (Grilli et al., 2018), external details were incidental (by instruction), and subtypes of external details were not assessed. We observed that ε4 carriers and non-carriers showed similar semantic production, whether it was for off-task semantic details in the AI (external semantic), on-task general semantic details produced in the GSAI, or personal semantic details produced in the PSAI (autobiographical fact, memories of repeated events, and self-knowledge). Thus, semantic types of autobiographical memory retrieval and general semantics appear to be relatively spared, and *APOE* ε4 carriers do not necessarily compensate for the subtle episodic memory impairment by producing additional personal or general semantic memory content. Previous research investigating the impact of *APOE* ε4 status on semantic memory performance via standard neuropsychological tasks indeed failed to report consistent impairment (Helkala et al., 1995; Laukka et al., 2013; Nilsson et al., 2006; Seidenberg et al., 2009; Wikgren et al., 2012), therefore concluding that the impact of *APOE* ε4 on semantic memory could be limited (Buckley et al., 2014; Wilson et al., 2002; for a review see Sacripante et al., 2025).

Taken together, these results suggest a robust reduction in episodic specificity linked to genetic risk for AD observed with multiple measures. Among healthy older adults with genetic risk for AD, autobiographical narratives therefore appeared less detailed relative to non-carriers. This could, arguably, represent a nuanced type of cognitive decline among carriers, reflecting subtle differences in narrative abilities whereby episodic specificity could be sensitive to ε4 status in cognitively unimpaired older adults (see Grilli et al., 2018).

Studies investigating changes in brain anatomy or connectivity indeed showed a link between *APOE* ε4 and changes in medial temporal lobe regions, including the hippocampus, that are known to be essential for episodic memory (Donix et al., 2010; Gallagher & Koh, 2011; Machulda et al., 2011; Mishra et al., 2018; for reviews see also Habib et al., 2017; Kucikova et al., 2021). Although the medial temporal lobes, in particular the hippocampus, appear to also be involved in certain semantic memory tasks (for a review see Duff et al. 2020), consistent with recent work that observed a neuroanatomical overlap between the semantic network and the episodic recollection memory (Binder & Desai, 2011; Irish et al., 2018; Renoult et al., 2019), they may not support all types of personal and general knowledge (Grilli & Verfallie, 2014; Martinelli et al., 2013; Renoult et al., 2012).

We note that in both the main analysis of target details (*Figure S1*) and the finer-grained analysis of detail categories (*Figure S2* and *Table S1*) in the *Supplementary Material*, the reported group effects reported for detail counts were no longer significant when proportions (e.g., internal-to-total) details were analysed. This is likely due to higher overall detail counts in non-carriers, reducing their proportional scores, although none of the detail categories other than episodic details under the standard AI instructions differed between groups.

Individuals with amnestic MCI and early AD also show a selective reduction in internal episodic details on the AI without the characteristic increase in external details seen in healthy aging (Simpson et al., 2023). The present results suggest that this dampening of age-related compensatory external detail production—along with reduced internal detail production—may be observed in individuals at genetic risk for AD even prior to clinical identification as in the case of amnestic MCI.

Finally, our results also suggest that healthy older adults retain the gist of their personal experience (Greene & Naveh-Benjamin, 2020, 2022, 2023, 2024; see also Sacripante et al., 2019, 2023), as memory for the gist of events is forgotten at a slower rate in time as compared to memory for more specific details (Brainerd & Reyna, 2015; Conway et al., 1991; Murphy & Shapiro, 1994; Reyna & Brainerd, 1995; Sachs, 1967; Sacripante et al., 2022; Sekeres et al., 2016; Thorndyke, 1977) and this could naturally make older adults’ autobiographical narratives more focused on gist and therefore accentuate the elaboration of semantic details over time (Lifanov et al., 2021; Spreng et al., 2018). In our study, the performance from the ε4 carriers’ group indeed suggests that the semanticization of their autobiographical narratives is robust and less sensitive to increased genetic risk for AD as opposed to episodic memory.

The present study however presents with some caveats. Firstly, the probed life chapters were not equally balanced across the three interview protocols, with participants being asked to recall specific events and personal life chapters from last year and ten years ago on the AI and PSAI, while general knowledge of culturally shared events was probed for last year only in the GSAI. However, scores in the AI and PSAI were averaged across time periods, so the difference should not be related to the number of time periods recalled. Furthermore, as noted by Melega et al. (2024), the GSAI was always administered at the end of the testing session, and this could have influenced the number of count details in this section of the interview, likely due to participants’ fatigue, which is not uncommon among healthy older adults, especially in prolonged interview protocols.

This study included only two homozygotes ε4ε4 carriers. The ε4 allelic variation is not particularly common in the general population (i.e., roughly 20-25%, Caselli & Reiman, 2012) and therefore even rarer for homozygotes ε4 (roughly 2% for European populations), though more of these participants would have allowed us to further explore whether the nuanced impairment in episodic memory could have been *APOE* ε4 dose-dependent, as demonstrated in some previous studies (Blacker et al., 1997; Davidson et al., 2006; Zimmermann & Butler, 2018). Lastly, our participants were not tested for any in vivo AD biomarkers (i.e., beta-amyloid or tau protein) so we could not verify whether any *APOE* ε4 carriers in our sample were effectively at risk for progression to AD, as specified by recent clinical guidelines (Dubois et al., 2021; see also Dubois et al., 2023).

## Conclusions

Episodic specificity is selectively reduced in healthy older with increased genetic risk of developing AD for on-task episodic details, as measured by well-established and novel interview protocols assessing autobiographical memory. These results were also corroborated by significantly lower performance in a test of word recognition memory among ε4 carriers. Nonetheless, carriers and non-carriers showed similar semantic memory production for on-task general and personal semantic details on the SAI as well as off-task semantic details in the AI.

Our findings are consistent with subtle differences in narrative abilities in healthy older ε4 carriers illustrated by a reduced production of episodic details, but a maintenance of the gist of their personal experience, perhaps through a process of semanticization that makes their semantic memories more robust and less sensitive to increased genetic risk for AD.

## Data Availability

Scored data and additional online materials are available at the Open Science Framework page of the project. Raw data cannot be provided as they cannot be fully anonymised.

https://osf.io/ewdzu/?view_only=8c57930ba2754594a0c6968c91403128

## Footnotes

1 In Melega et al. (2024) general semantic testing was restricted to last year only in the context of a comparison between younger and older adults. Pilot data showed that younger participants had significant difficulties in recalling public events and culturally shared knowledge from ten years before the interview (e.g., childhood times).

2 Post-hoc tests revealed that carriers’ average scores in Autobiographical Facts were significantly higher than non-carriers (*p* = .022). It is however likely that this significant difference was masked after using a seven-level factor (Detail Type).

