## Supplementary Material for "Assessing Episodic and Semantic Autobiographical Recall in Healthy Older *APOE* ε4 Carriers"

**Supplementary materials**

#### **Section 1. DNA extraction and *APOE* Genotyping**

DNA was collected via sterile brush or cotton buccal swabs. To ensure participants had not had any food or drink 30 minutes prior to the swab, this step was always completed after the task. All swabs were air-dried at least two hours before processing and all swabs were processed within one week. DNA was extracted according to the manufacturer’s instructions using the Qiagen Buccal Swab Spin protocol (QIAamp DNA MiniKits, Qiagen, Hilden, Germany). AE buffer was used for the final elution step. In brief, the tips of the buccal swabs were combined with PBS, QIAGAN Protease stock and AL buffer and incubated at 56 ˚C for ten minutes. After this ethanol was added and the solution added to a QIAmp Mini spin collum and centrifuged. In subsequent steps Buffer AW1 and AW2 were added to the QIAmp Mini spin column and centrifuged. The recommended Step 9 where the QIAmp Mini spin collum is centrifuged again without additional buffer to avoid carryover was included. In a last step, DNA was eluted in Buffer AE.

DNA yield was measured through absorbance ratios (260/230 and 260/280) using the NanoDrop (NanoDrop ™ 1000, Thermo Fisher Scientific, Waltham, MA, USA) and stored at -30˚C until quantitative Polymerase Chain Reaction (qPCR) was performed. Genomic DNA was isolated and purified using the QIAmp DNA Mini Kit (QIAGEN). Extracted DNA was then quantified by a NanoDrop™ spectrophotometer (Thermo Fisher Scientific). Single-Nucleotide Polymorphism (SNP) *APOE* genotyping (rs429358 and rs7412) was performed using TaqMan® Predesigned SNP Genotyping Assays (Assay ID: C___3084793_20 and C____904973_10, Thermo Fisher Scientific) following manufacturer’s instruction. Briefly, for each reaction, 2µL of DNA template containing 1–20 ng extracted DNA, 0.5µL of 20x Assay Working Stock, 5µL of 2x qPCRBIO Probe Mix Lo-ROX (PCR Biosystems), and 2.5µL UltraPure™ nuclease-free water (Thermo Fisher Scientific) were mixed to make up a total volume of 10μL. Real-time qPCR and post-PCR analysis were carried out in an ABI 7500 Fast Real-Time PCR System (Applied Biosystem, Thermo Fisher Scientific).

### **Section 2. The effect of carriers’ status on internal and external details in the AI**

To test the impact of APOE ε4 status on internal and external details produced in the AI as in Grilli et al. (2018), we computed a mixed 2x2 ANOVA with Group as between-subjects variable (Carrier vs Non-carrier) and Detail Type as within-subjects variable (Internal details vs External details). *Figure S1* illustrates the cumulative count scores of internal and external details produced by the two groups in the original AI.

ANOVA revealed a main effect of Detail Type, *F*(1,72) = 14.60, η2p = 0.17, 95% CI [0.06, 1.00], p < .0001, as participants in both groups produced a higher amount of internal details (M = 77.75, SD = 27.01) as compared to external details (M = 53.95, SD = 29.69). There was also a significant main effect of Group, F(1,72) = 8.949, η2p = 0.11, 95% CI [0.02, 1.00], p < .01, since Carriers overall produced a lower amount of details (*M* = 56.53, *SD* = 31.62) than Non-carriers (*M* = 75.16, *SD* = 26.90), *t*(72) = 2.992, *p* < .01. However, the interaction between Detail Type and Group was non-significant, F(1,72) = 0.011, η2p = 0.00, 95% CI [0.00, 1.00], p = 0.91. This analysis thus suggests that carriers produced less internal and external details than non-carriers. For external details, this is consistent with the general trend of a small decrease in all subtypes of externals in carriers compared to non-carriers in the AI (see *Table 3* in main manuscript), though group differences did not approach significance for any of these subtypes of externals taken individually (see main manuscript, *APOE group differences in the production of target details across interviews*).

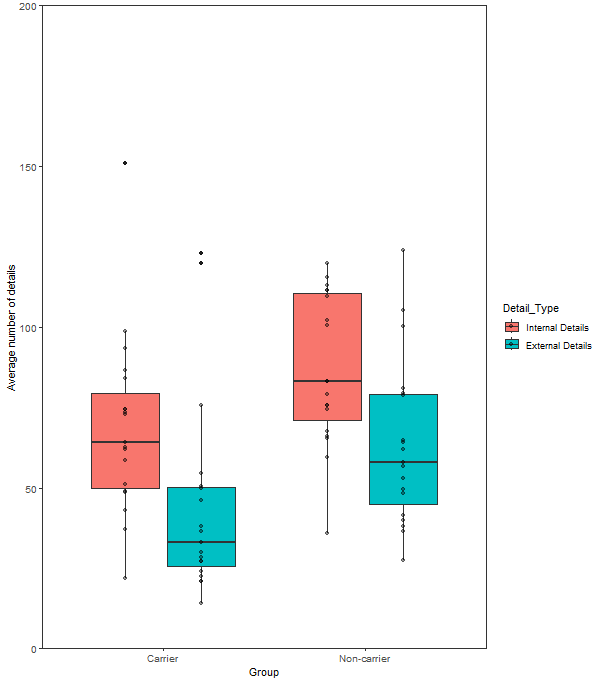

*Figure S1*. Boxplots showing counts of Internal and External Details on the original Autobiographical Interview in the Carrier and Non-carrier groups.

### **Section 3. *APOE* group differences in the production of target details across interviews**

We also ran an analysis with proportional scores, to correct for overall output differences (i.e., verbosity of participants). *Figure S2* illustrates the cumulative proportional target details produced by carriers and non-carriers in each interview (for descriptive statistics see *Table S1)*. As with count scores, there was a main effect of Interview, *F*(2,108) = 80.422, *η2_p_* = 0.60, 95% CI [0.50, 1.00], *p* < .0001, where participants in both groups produced the highest proportion of details on the PSAI (*M* = 0.76, *SD* = 0.06) as compared to the AI (*M* = 0.68, *SD* = 0.09) and GSAI (*M* = 0.44, *SD* = 0.15). However, the main effect of Group was non-significant, *F*(1,108) = 0.273, *η2_p_* = 0.002, 95% CI [0.00, 1.00], *p =* 0.603, indicating that carriers and non-carriers overall generated similar proportions of target details across interviews. The interaction between Group and Interview type was also non-significant, *F*(2,108) = 1.566, *η2_p_* = 0.02, 95% CI [0.00, 1.00], *p =* 0.092.

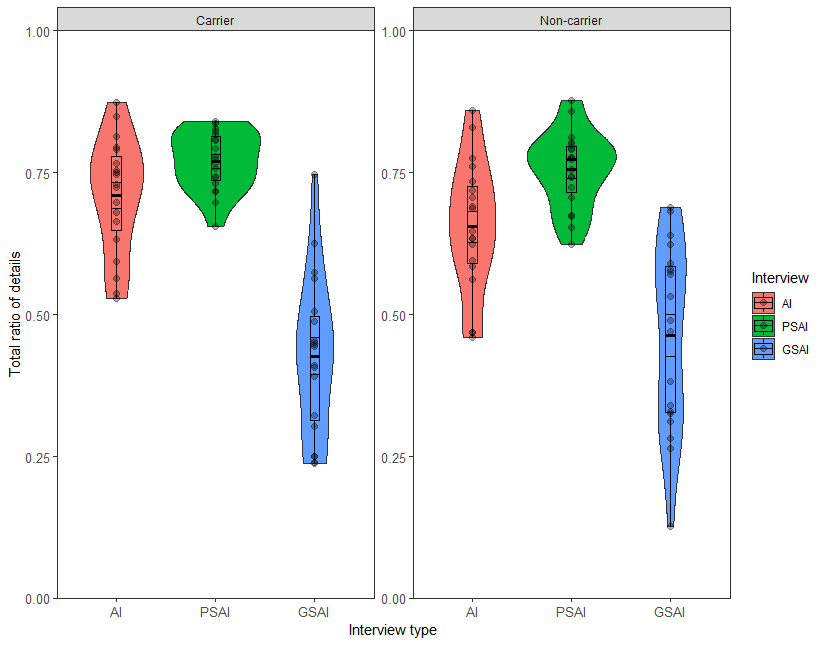

*Figure S2.* Proportions of target detail in cumulative recall in the carrier and non-carrier groups across the Autobiographical Interview (AI), the Personal Semantic Interview (PSAI) and the General Semantic Interview (GSAI), with medians and interquartile range.

### **Section 4. APOE group differences in detail elaboration in each interview**

As for count details, we here also provide a fine-grain analysis of the production of detail categories across the three interviews with proportional scores (see *Figure S3*). Average proportional scores on each detail category can be found in *Table S1*.

##### **Autobiographical Interview**

Average proportional scores on the original AI are illustrated in *Figure S3* (top panel). There was a significant main effect of Detail Type, *F*(6,288) = 702.36, *η2_p_* = 0.94, 95% CI [0.93, 1.00], *p* < .0001). Post-hoc pairwise tests revealed that the overall group of participants produced a greater proportion of Episodic details as compared to all other detail categories (Autobiographical Facts: p < .0001, Self-Knowledge: p < .0001, Repeated Events: p < .0001, General Semantic: p < .0001, Repetitions: p < .0001, Other: p < .0001).

The main effect of group was not significant, *F*(1,288) =0.00, *η2_p_* = 0.00, 95% CI [0.00,1.00], *p* = 1.00. Both carriers and non-carriers overall produced a similar proportion of episodic details, as seen by the lack of interaction between Detail Type and Group, *F*(6,288) = 1.651, *η2_p_* = 0.04, 95% CI [0.00, 1.00], *p =* .134.

##### **Personal Semantic Interview**

When analysing the proportional data of the subtypes of details produced in the PSAI (see *Figure S3*, middle panel), there was again only a main effect of Detail Type, *F*(6,252) = 388.104, *η2_p_* = 0.90, 95% CI [0.89, 1.00], *p* < .0001). Post-hoc pairwise tests showed that the overall group of participants produced a greater proportion of Autobiographical Facts as compared to any other category of details (Episodic details: p < .0001, Self-Knowledge: p < .0001, Repeated Events: p < .0001, General Semantic: p < .0001, Repetitions: p < .0001, Other: p < .0001). Also, the overall group produced a higher proportion of Self-Knowledge details compared to any other detail categories (Episodic details: p < .0001, Repeated Events: p < .0001, General Semantic: p < .0001, Repetitions: p < .0001, Other: p < .0001) except for Autobiographical Facts. Finally, participants produced a greater proportion of Repeated Events details than Episodic details (p < .0001), General Semantic (p < .0001) and Repetitions (p <.0001). These findings suggest that participants were overall on task, as seen by their proportional scores in target details such as Autobiographical Facts, Self-Knowledge and Repeated Events.

The main effect of group was non-significant, *F*(1,252) = 0.000, *η2_p_* = 0.00, 95% CI [0.00, 1.00], *p* = 1.00, as well as the interaction between Detail Type and Group, *F*(6,252) = 0.344, *η2_p_* = 0.00, 95% CI [0.00, 1.00], *p* = .918. This indicates that carriers and non-carriers were equally on task in their semantic production for personally relevant events.

##### **General Semantic Interview**

Average proportional scores on the different subtypes of details are illustrated in *Figure S3* (bottom panel)*.* Once again, there was a main effect on Detail Type, *F*(6,252) = 110.609, *η2_p_* = 0.72, 95% CI [0.68, 1.00], *p* < .0001. Post-hoc pairwise tests were carried out and revealed that the overall group of participants produced a greater proportion of general semantic details as compared to any other detail categories (Episodic details: p < .0001, Autobiographical Facts: p < .0001, Self-Knowledge: p < .0001, Repeated Events: p < .0001, General Semantic: p < .0001, Repetitions: p < .0001, Other: p < .0001). Interestingly, participants also produced a greater proportion of Self-Knowledge details when compared to all the other detail categories (Episodic details, p < .001, Autobiographical Facts: p < .0001, Repeated Events: p < .0001, Repetitions: p < .0001, Other: p < .001), apart from general semantic details. These comparisons indicate that participants were overall on task, given the highest scores on target General Semantic details.

The main effect of Group was non-significant, *F*(1,252) = 0.00, *η2_p_* = 0.00, 95% CI [0.00, 1.00], *p* = 1.00, as well as the interaction between Detail Type and Group, *F*(6,252) = 0.037, *η2_p_* = 0.02, 95% CI [0.00, 1.00], *p* = 0.643, which indicates that participants from both groups were on-task in the production on internal general semantic details.

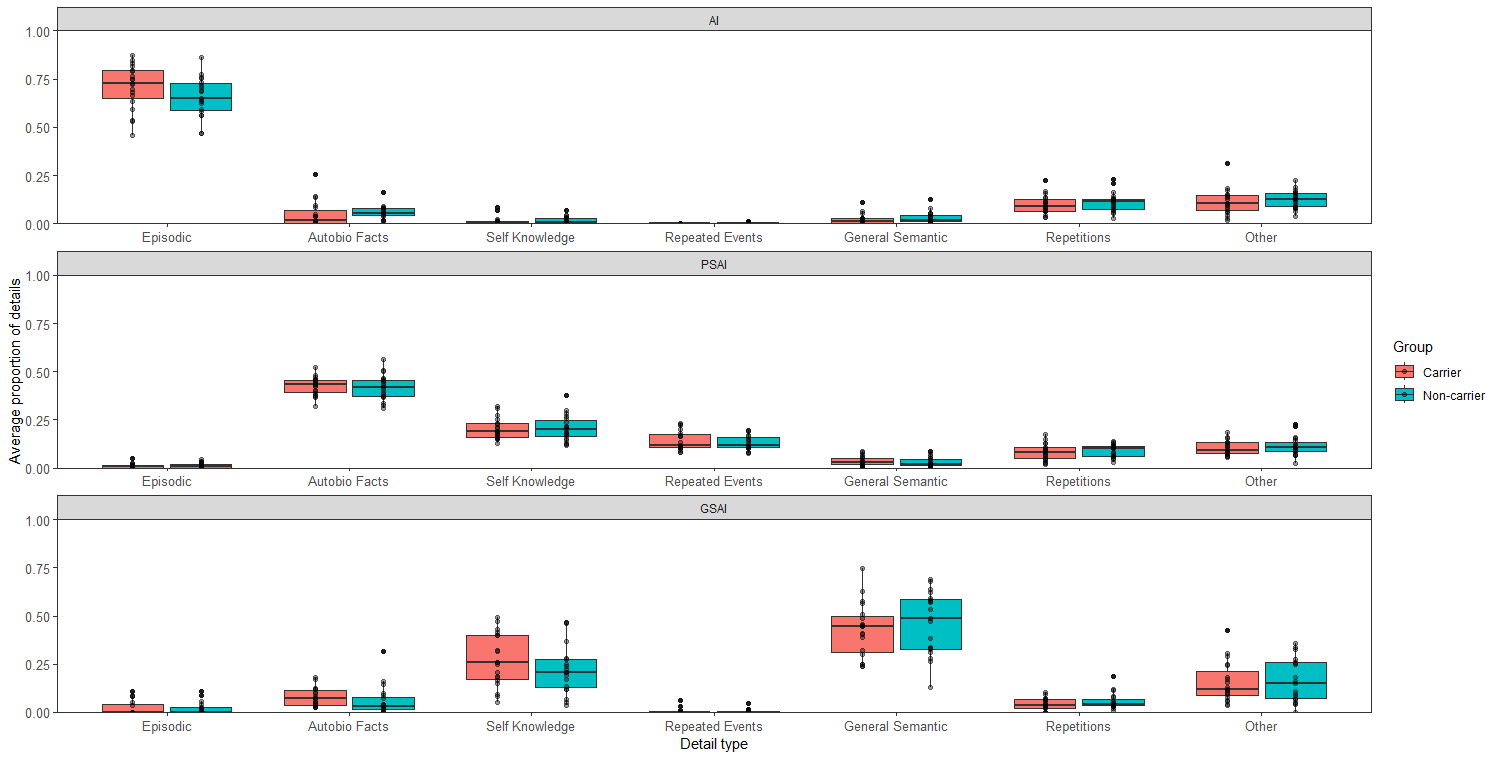

*Figure S3*. Proportions of detail types during cumulative recall in the Carrier and Non-carrier group in the original Autobiographical Interview (AI), Personal Semantic Interview (PSAI), and General Semantic Interview (GSAI).

| ***Table S1*. Proportional scores in Carriers and Non- carriers for cumulative recall (Free Recall, General Probe, and Specific Probe) in the AI, PSAI, and GSAI** | | | | | | |
| --- | --- | --- | --- | --- | --- | --- |
|  | AI | | PSAI | | GSAI | |
| Detail type | Carriers | Non-carriers | Carriers | Non-carriers | Carriers | Non-carriers |
| Episodic | **0.71 (0.11)** | **0.66 (0.10)** | 0.01 (0.01) | 0.01 (0.01) | 0.02 (0.03) | 0.02 (0.03) |
| Autobio Fact | 0.05 (0.07) | 0.06 (0.03) | **0.43 (0.04)** | **0.42 (0.06)** | 0.08 (0.04) | 0.06 (0.08) |
| Self-knowledge | 0.01 (0.02) | 0.01 (0.02) | **0.20 (0.05)** | **0.21 (0.06)** | 0.27 (0.13) | 0.23 (0.13) |
| Repeated Events | 0.00 (0.00) | 0.00 (0.00) | **0.14 (0.05)** | **0.13 (0.03)** | 0.00 (0.01) | 0.00 (0.01) |
| General Semantic | 0.02 (0.02) | 0.03 (0.03) | 0.04 (0.02) | 0.03 (0.02) | **0.43 (0.14)** | **0.47 (0.16)** |
| Repetitions | 0.10 (0.05) | 0.11 (0.05) | 0.08 (0.04) | 0.09 (0.03) | 0.04 (0.03) | 0.06 (0.04) |
| Other | 0.11 (0.06) | 0.13 (0.05) | 0.10 (0.03) | 0.11 (0.05) | 0.16 (0.10) | 0.16 (0.11) |
